# A one-step deep learning framework for psoriasis area and severity prediction trained on interventional clinical trial images

**DOI:** 10.1101/2023.03.23.23287628

**Authors:** Yunzhao Xing, Sheng Zhong, Samuel L. Aronson, Francisco M. Rausa, Dan E. Webster, Michelle H. Crouthamel, Li Wang

## Abstract

Image-based machine learning holds great promise for facilitating clinical care, however the datasets often used for model training differ from the interventional clinical trial-based findings frequently used to inform treatment guidelines. Here, we draw on longitudinal imaging of psoriasis patients undergoing treatment in the Ultima 2 clinical trial (NCT02684357), including 2,700 body images with psoriasis area severity index (PASI) annotations by uniformly trained dermatologists. An image-processing workflow integrating clinical photos of multiple body regions into one model pipeline was developed, which we refer to as the ‘One-Step PASI’ framework due to its simultaneous body detection, lesion detection, and lesion severity classification. Group-stratified cross-validation was performed with 145 deep convolutional neural network models combined in an ensemble learning architecture. The highest-performing model demonstrated a mean absolute error of 3.3, Lin’s concordance correlation coefficient of 0.86, and Pearson correlation coefficient of 0.90 across a wide range of PASI scores comprising disease classifications of clear skin, mild, and moderate-to-severe disease. Within-person, time-series analysis of model performance demonstrated that PASI predictions closely tracked the trajectory of physician scores from severe to clear skin without systematically over or underestimating PASI scores or percent changes from baseline. This study demonstrates the potential of image processing and deep learning to translate otherwise inaccessible clinical trial data into accurate, extensible machine learning models to assess therapeutic efficacy.

## Introduction

Psoriasis is a chronic inflammatory disease that damages the skin and impacts several other organ systems.^1^ Psoriasis is driven by a confluence of genetic, immunologic, and behavioral factors and affects between 2-4% of the population worldwide with overall prevalence across demographic groups.^2^ While effective systemic and targeted therapies have been developed for psoriasis, non-treatment and undertreatment remain significant problems, with over 50% of patients reporting dissatisfaction with their treatment.^3^

Consensus treatment guidelines for psoriasis use the Psoriasis Area Severity Index (PASI) clinical assessment to measure disease severity and provide target values for treatment efficacy.^4^ The PASI assessment is performed by a trained physician evaluating the overall body surface area (BSA) or involvement that is affected by psoriatic plaques and assessing the severity of plaques in the categories of erythema (redness), induration (thickness), and desquamation (scaling) to generate a composite score on a scale of 0–72.^5^ The PASI score and thresholds for percent change upon treatment (i.e. PASI75, PASI100) are routinely used for eligibility criteria and primary endpoints of therapeutic efficacy in interventional clinical trials.

Image-based machine learning workflows to generate PASI scores have potential future use to ease the burden of PASI assessment in clinical trial, clinical care, and remote monitoring applications.^6^ Prior studies have employed image-based machine learning models to perform PASI assessments.^7–10^ However, these efforts have not trained their machine learning models on interventional studies collecting longitudinal, standardized imaging data. As a result, machine learning assisted PASI scoring may not accurately measure individual’s change upon treatment, potentially hindering deep learning adoption to assess therapeutic efficacy in clinical trial and routine clinical practice settings.

Here, we have used interventional clinical imaging data to train a deep learning workflow, called the ‘one-step PASI’ framework, to integrate clinical photos of multiple body regions into one model pipeline for simultaneous body detection, lesion detection, and lesion severity classification to generate PASI scores.

## Results

### Dataset

To develop a deep learning workflow for prediction of PASI scores from imaging data, we utilized 2700 standardized photos captured during UltIMMa-2: a randomized, placebo-controlled phase 3 interventional trial (NCT02684357) investigating the efficacy of Risankizumab to reduce the severity of psoriasis over the course of 16 weeks.^11,12^ 74.8% of study participants achieved 90% reduction in PASI score (PASI90) during the first 16 weeks, providing a large dynamic range of PASI scores within individuals.

This imaging dataset included 60 psoriasis patients with images captured for five or six site visits totaling 338 visits across all participants. At each visit, site staff used a Nikon D5100 camera to capture 4928×3264 pixel photos of three key body regions: four photos of upper extremities, two of the trunk, and two of the lower extremities (Supplemental Figure 1). Head and neck photos were not collected to preserve subject privacy. The three regions were scored in person in a clinical setting by uniformly trained dermatologists for region involvement, erythema, induration, and scaling severity.

To balance the training and testing data sets, images were sorted into training or testing datasets with a ratio of 90% training and 10% testing. Images associated with an individual subject were all assigned to either the training or testing set, ruling out potential within-subject information leakage between training and test sets. The training and testing datasets contained images with a similar distribution of erythema, induration, scaling, and involvement (Figure 2).

### Image pre-processing and tiling

To enable the workflow to read a variable number of images from the three different body regions in a standardized way, 8 raw images per patient were resized to 128×128 pixels and tiled together in 2×2 grid pattern to form a single composite image (Figure 1). Solid black squares filled in missing regions of the 2×2 composite image grids generated for trunk and lower extremity regions because they each had two images. The four upper extremity images completed the 2×2 composite. Black squares contained no features for models to detect, and therefore did not interfere with learning. Sub-images comprising the 2×2 composite were randomly rotated and their positions within the grid were shuffled to avoid position-detection bias in the trained PASI scoring model. The output of pre-processing was a set of three 2×2 composite images, one for each body region, for each clinical visit. Images were also pre-processed with randomized brightness and contrast adjustments. Images used for training and testing were in full color, but are converted to black and white for demonstration purposes and privacy protection.

**Figure 1.**
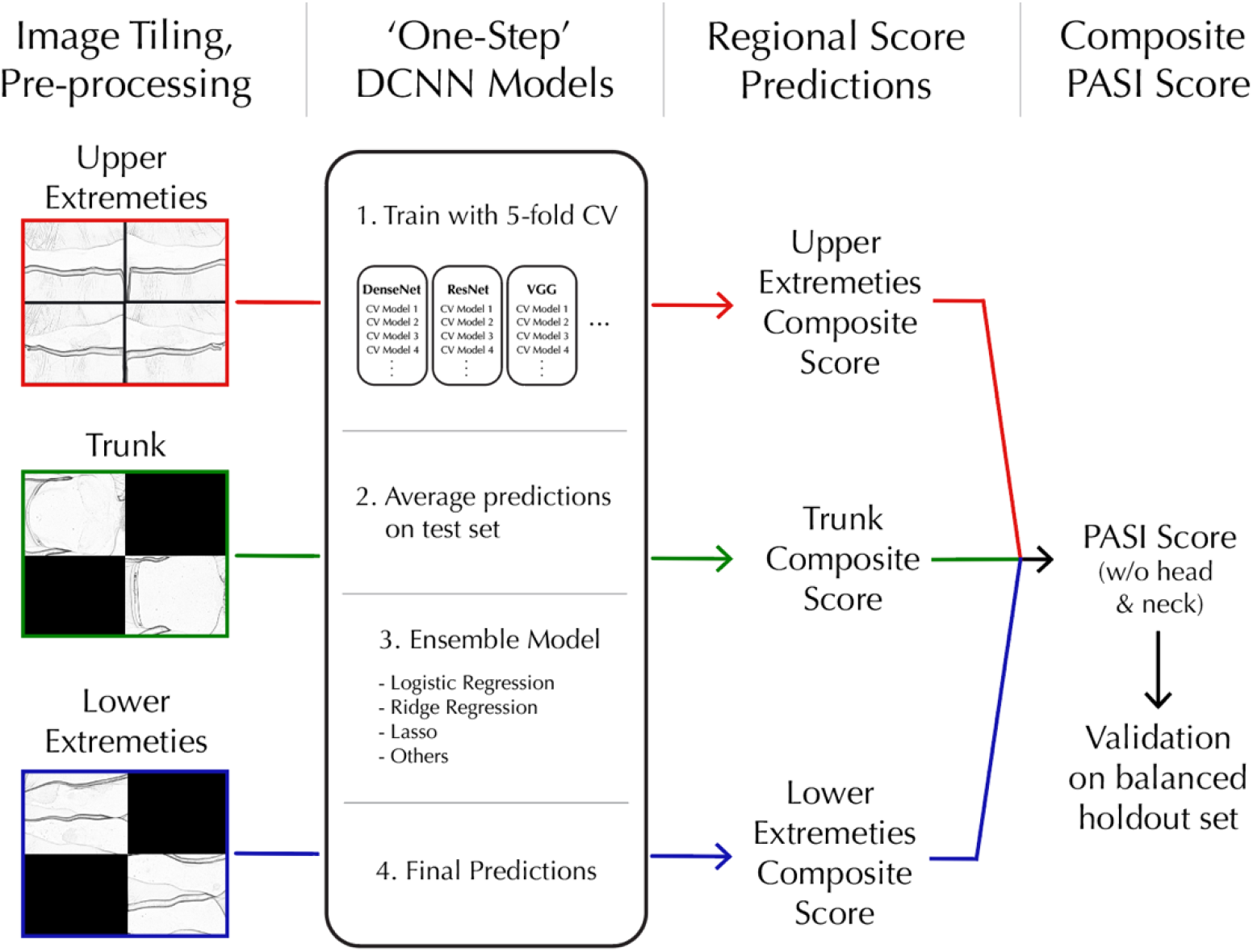
One-step workflow for processing whole-body imaging, training DCNN and evaluation of composite PASI scores.

### Model training and performance

Both base and meta learners were validated for PASI scoring accuracy using the test dataset. The mean predictions from the five deep convolutional neural networks (DCNNs) in each base-learner were used for testing. Learning models were evaluated using mean absolute error (MAE), mean squared error (MSE), Lin’s concordance score (CCC), and person correlation coefficient (PCC) (Table 1).

**Table 1.** Top 10 models based on MAE performance. (MAE=mean absolute error; PCC=pearson’s correlation coefficient; MSE= mean squared error; CCC=Lin’s concordance correlation coefficient)

| Model | MAE | PCC | MSE | CCC |
| --- | --- | --- | --- | --- |
| **resnet34** | **3.336** | **0.9** | **27.191** | 0.83 |
| shufflenet_v2_x0_5 | 3.479 | 0.896 | 23.616 | **0.864** |
| densenet121 | 3.507 | 0.892 | 27.475 | 0.835 |
| resnet152 | 3.539 | 0.885 | 28.499 | 0.825 |
| resnet101 | 3.639 | 0.866 | 28.821 | 0.835 |
| vgg19_bn | 3.676 | 0.87 | 30.606 | 0.826 |
| resnext50_32x4d | 3.709 | 0.883 | 30.273 | 0.812 |
| meta_learner_ridge | 3.781 | 0.879 | 30.583 | 0.812 |
| mobilenet_v2 | 3.786 | 0.886 | 29.025 | 0.823 |
| vgg11_bn | 3.788 | 0.874 | 31.482 | 0.806 |

The resnet34 model.^13^ demonstrated the best performance as measured by MAE, MSE, and PCC in DCNN validation testing. The base-learner models largely outperformed the four meta-learning architectures. The best meta-learner was the logistical regression model with ridge regularization, but was ranked 8^th^ among all learners.

To explore the model’s performance in the test dataset, ground-truth physician PASI scores were plotted with the resnet34 PASI score predictions comprising 34 clinical trial visits across 6 subjects (Figure 3). There was a strong correlation (p=0.90) across the range of ground-truth PASI scores between 0 and 43.7, a range that covers overall psoriasis severity classifications of clear, mild, and moderate-to-severe disease.^14^ To explore the one-step workflow for its capacity to track changes in an individual’s psoriasis severity over time, time-series analysis of individuals over the course of the study was performed, revealing that one-step PASI estimations closely tracked the trajectory of the physician-based PASI scores upon interventional treatment. The best performing model did not systematically over- or underestimate PASI scores or the interventional response variable that denotes percent change from baseline (i.e. PASI75) (Figure 4).

**Figure 2.**
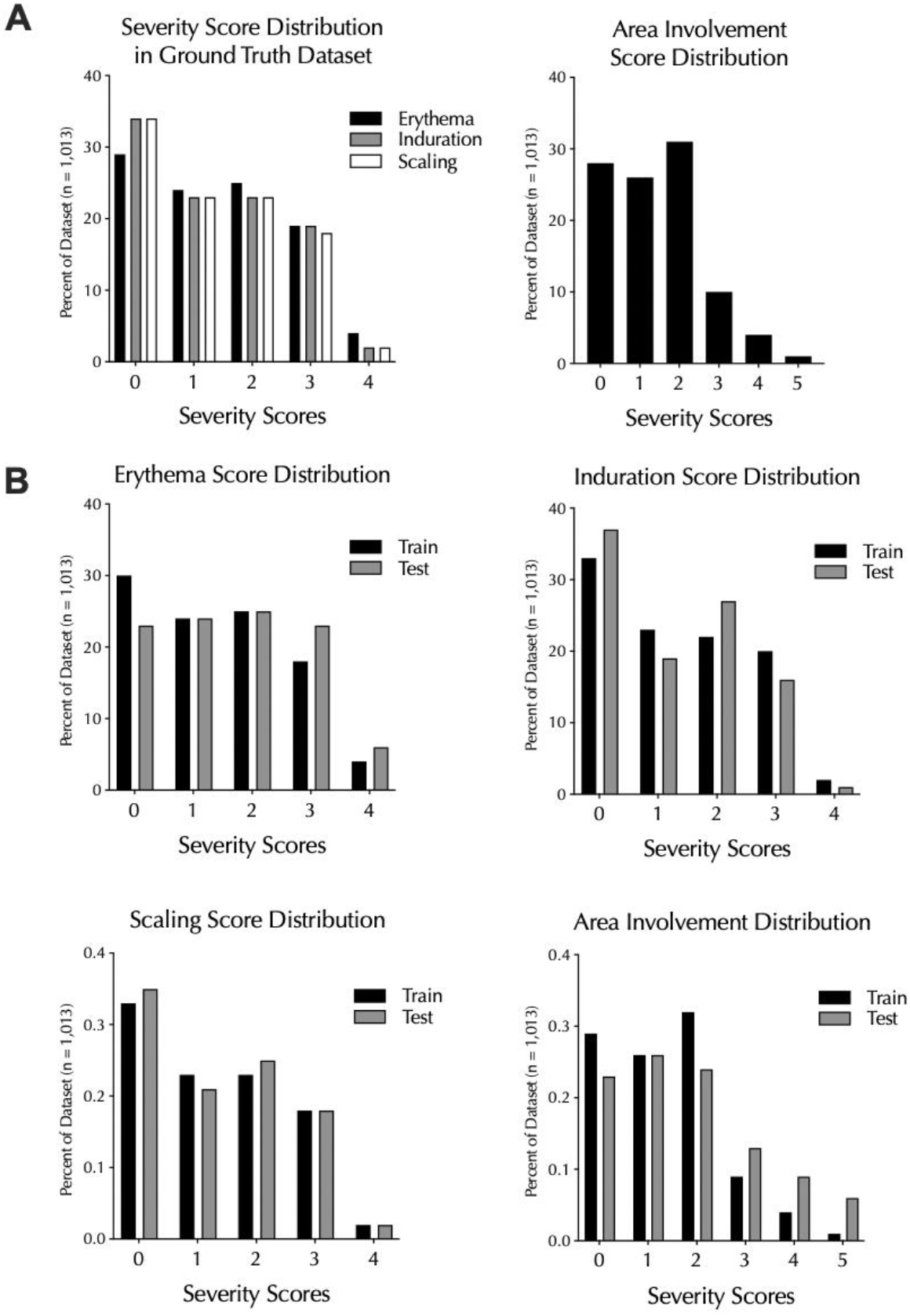
Distribution of Psoriasis severity scores and involvement (or body surface area) scores in (A) the ground truth annotated dataset overall and (B) their distribution between training and test sets.

**Figure 3.**
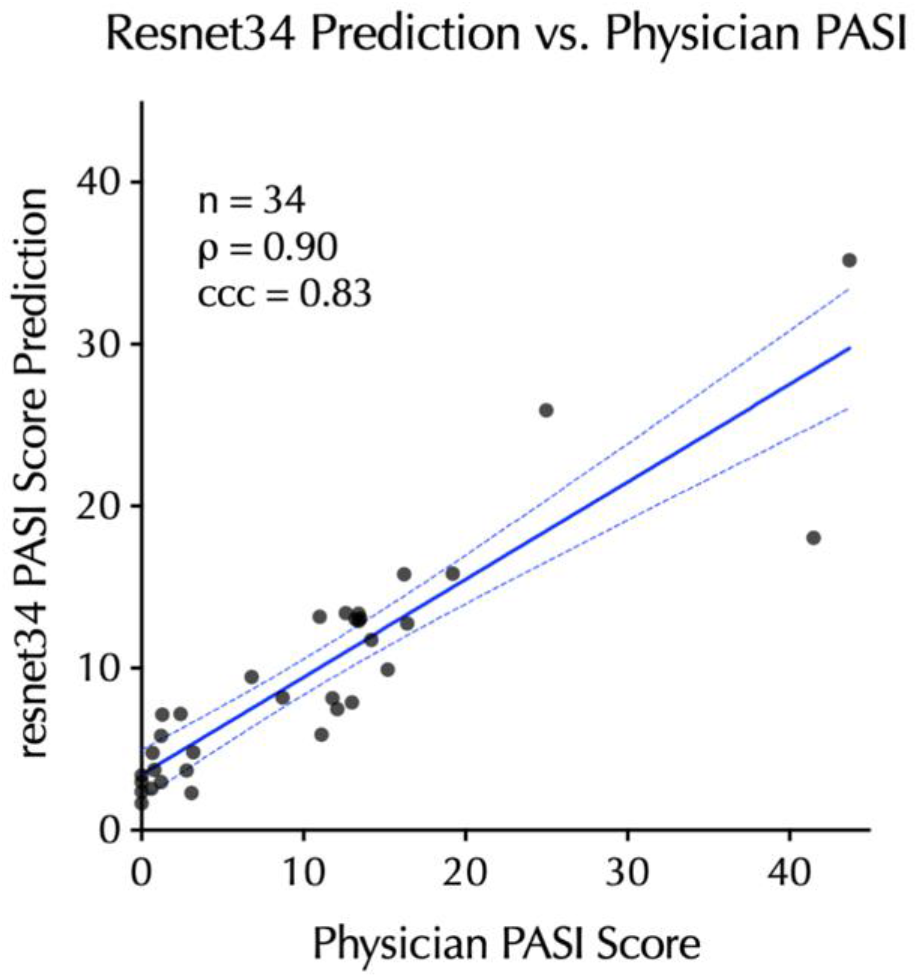
Correlation between the top-performing PASI estimation model (resnet34) predictions on the test dataset relative to in-clinic physician PASI scores. 95% CI is shown as dashed-blue lines.

**Figure 4.**
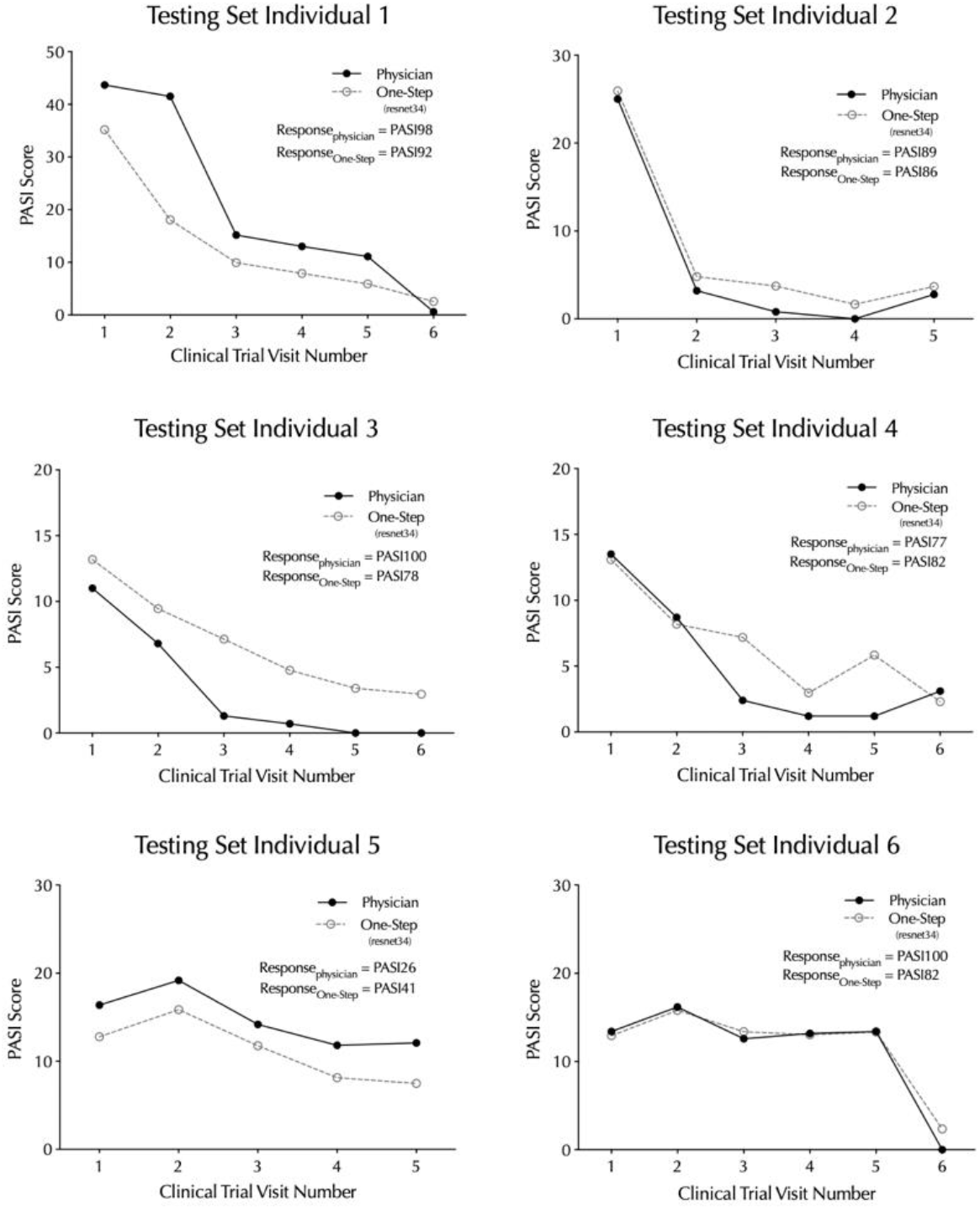
Time-series of top-performing One-Step PASI estimation model (resnet34) predictions relative to in-clinic physician PASI scores. The clinical trial endpoint metric, percent change from baseline in the PASI score (e.g. PASI98, PASI92), is listed for the model and physician.

## Discussion

In this study, we developed a ‘one-step’ deep learning framework for multi-image processing and deep convolutional neural network (DCNN) training to predict psoriasis area severity index (PASI) scores. This framework utilized 2700 images of psoriasis patients’ skin while undergoing treatment in a controlled clinical setting to train and test model performance. The best-performing model trained by the framework (resnet34) produced PASI predictions demonstrating a strong correlation (Pearson’s r = 0.9) with in-person physician scoring and a mean absolute error of 3.3 on the PASI scale of 0 to 72. Importantly, the PASI score estimations for individuals over time tracked the trajectory of physician scoring, without systemic over- or underestimates of PASI scores at an individual visit or the estimated percent change over time that is typically used in clinical trial settings (e.g. PASI75).

A key component of models from the one-step PASI framework is their training on highly standardized, interventional clinical trial data. In contrast to deep learning workflows trained with a large volume of cross-sectional images capturing a snapshot in time from a given patient, the UltIMMa-2 trial images allow the model to learn in the setting of within-person disease changes from baseline over time. In this dataset, individuals often progressed from severe disease to clear skin. Thus, in addition to evaluating single-point diagnostic accuracy, we investigated one-step models for their capacity to accurately assess therapeutic efficacy.

PASI scoring requires three different tasks: body detection from background, disease lesion segmentation from healthy skin, and severity classification for detected lesions. Other methods use separate DCNNs to achieve each of the three tasks. For example, previous work from Li et al. used 86,000 images labeled with lesion location for the segmentation. The one-step PASI framework implicitly integrates all three tasks into one DCNN model by pre-processing images from multiple body regions into a standard input. This allows the model to generalize learning from a relatively small sample size by connecting information across multiple body regions. Compared to a benchmark model ResNet-50,^10^ the resnet34 model trained by the one-step PASI framework demonstrated better MAE (3.34 vs 3.50) while also training on fewer images (2700 vs. 5205).

Despite the strong concordance between the one-step PASI predictions and ground truth PASI scores, there are limitations of this work. The dataset contained no head and neck region photos. While there are regional severity scores for other body regions, the accuracy of facial, head, and neck region scoring is unknown. The imaging to generate the dataset was highly standardized so it is unknown how these models would perform with non-clinical image capture methods such as a smartphone.

There was a relatively small sample size used to train the DCNN model. Access to a larger dataset of clinical trial imaging would enable more robust DCNN training and allow for assessment of how well these models generalize to a larger population. This is particularly important for clinical applications in dermatology, where imbalances in skin tone representation in the training set may impart systematic biases in digital tools.^15,16^ We envision this method to be employed on larger datasets in the future to evaluate this important limitation.

In summary, the one-step PASI deep learning framework trains models can accurately estimate PASI scores from clinical imaging. While further work with larger datasets will be needed to generalize this approach, this study demonstrates the potential of image processing and deep learning to translate otherwise inaccessible clinical trial data into accurate, extensible machine learning models with the potential to assess therapeutic efficacy.

## Methods

### Data Pre-processing

For each subject-visit, 8 raw images were collected: four images of upper extremities, two of the lower extremities, and two of the trunk. Images were resized from the raw 4928*3264 pixels to 128*128 pixels. Despite pixel reduction, the resized images maintained original ratios to prevent distortion of relative involvement scores used for PASI scoring.

For each of the three body regions, resized photos were stitched together in 2×2 grid pattern to form a single composite image. Solid black squares filled in missing regions of the 2×2 composite image grids generated for trunk and lower extremity regions because they each had two images. The four upper extremity images completed the 2×2 composite. Black squares contained no features for DCNN models to detect, and therefore did not interfere with learning. Sub-images comprising the 2×2 composite were randomly rotated and their positions within the grid were shuffled to avoid position-detection bias in the trained PASI scoring model. The output of pre-processing was a set of three 2×2 composite images, one for each body region, for each clinical visit.

Images were also pre-processed with randomized brightness and contrast adjustments. Images used for training and testing were in full color but were converted to black and white for demonstration purposes in this manuscript to protect subject privacy.

### Train/Test Splitting

A group-stratified train-test splitting strategy was applied based on a greedy algorithm. Subject records were randomly shuffled into groups in a stepwise fashion, where train or test group assignments were made one subject at a time. This minimized scoring imbalances by assessing score distributions after each round of sorting. The scaling severity score was used to guide data splitting.

Scaling (desquamation) was the chosen guiding metric for data splitting because it has the highest correlation with other severity dimensions. Our imbalance measure is the average of absolute train-test differences in the adjusted percentage out of the total number of records for a given desquamation severity label over five labels, where the adjustment accounts for the different sizes of the cohorts. To generate the most representative dataset split, random shuffling was done multiple times with different random seeds.

### PASI Scoring Algorithm

PASI scores are generated by 1) scoring the disease severity of multiple body regions and 2) combining all regional severity scores into a weighted composite PASI score for the whole body.

Disease severity is determined by evaluating body regions for the degree of erythema, induration, scaling (i.e., desquamation), and lesion body surface area (BSA) (i.e., involvement). A regional composite score is calculated as seen in Equation 1 where *E*_*region*_ *I*_*region*_, and *S*_*region*_ are the erythema, induration, and scaling severity and *B*_*region*_ is the BSA score for psoriasis lesions.

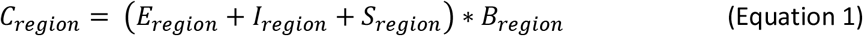

Next, a PASI score is calculated based on a linear combination of the regional composite scores of all four individual regions.^5^ Due to subject privacy protection, there are no head and neck photos in the dataset used in this paper. A PASI sub-score without head and neck disease severity is derived using Equation 2. *C*_*upper*_, *C*_*trunk*_, and *C*_*lower*_ are the regional composite score of upper extremities, trunks, and lower extremities specifically.

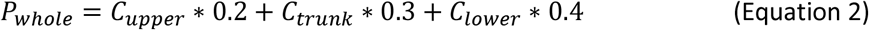

### Learning Model Architecture

The one-step workflow trained a DCNN module with an ensemble learning architecture using the prespecified training dataset. This included base-learners with 5-fold cross-validation. 29 pre-trained DCNN models were imported from PyTorch packages for use as base learners.^17^ The complete list of base learner models is available in Supplemental table 1.^13,18–29^ For each of the 29 base-learner DCNN architectures, 5 DCNN copies were created. Each of the DCNN copies was fine-tuned with 4 folds (out of 5 total folds) from the training dataset and subsequently predicts the last fold (1 out of 5 folds). 145 DCNNs were used across the 29 base learners. Predictions for each sample in the training dataset were generated by base-learners and concatenated into one vector input to train the ensemble model. Four different logistic regression models were used for the ensemble model. These included logistic regression (meta_learner), logistic regression with intercept term (meta_learner_w_intercept), lgosistic regression with ridge penalty (meta_learner_ridge), and ridge penalty (meta_learner_ridge_w_intercept).

The one-step workflow tested the trained model by having each of the 5 DCNN models within the 29 base-learners make predictions for the full testing dataset. For each image, the mean of the 5 predictions generated by each base-learner was calculated. The 29 mean predictions generated for each image were concatenated into a vector input to the ensemble model. Finally, the ensemble model predicted the PASI score results for the testing dataset.

### Implementation

A Dell Precision 7550 laptop with an Intel i7-10850H CPU and Nvidia Quadro TRX 4000 graphic card was used for this workflow. The pre-trained DCNNs were imported from PyTorch (v.1.9.0),^17^ with classification layers being updated for regression tasks. The training and testing pipeline was implemented with PyTorch and PyTorch Lightning (v1.4.0).^30^ Mean squared error was the loss criterion and the optimizer was the stochastic gradient descent (SGD) with momentum set to 0.9. The learning rate was initialized as 0.01 and reduced by 0.1 at the plateau. The plateau threshold was set to 0.0001, and the patience was 10. The training batch size was 16. Early stopping was applied during the training procedure with patience of 50 epochs.

## Data Availability

All image and annotation data used in this work is prohibited from broader distribution due to the data privacy outlined in the informed consent obtained from clinical trial subjects.

## Supplemental Data

**Figure S1.**
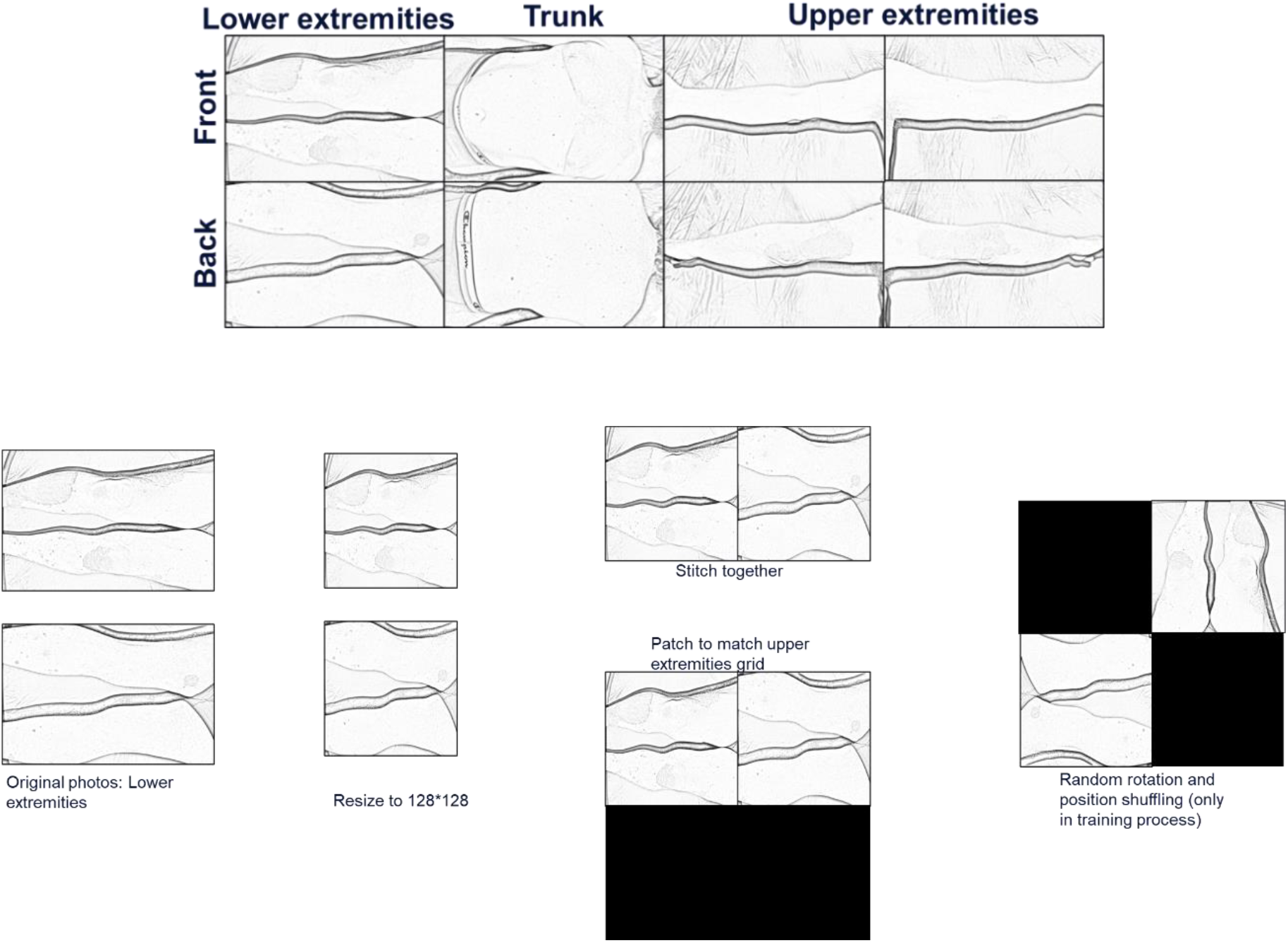
Illustration of eight clinic photos for each visit and the imaging preprocessing steps. The images are transformed to grayscale for privacy consideration only. The original full-color images are used to train the models.

**Table S1:**
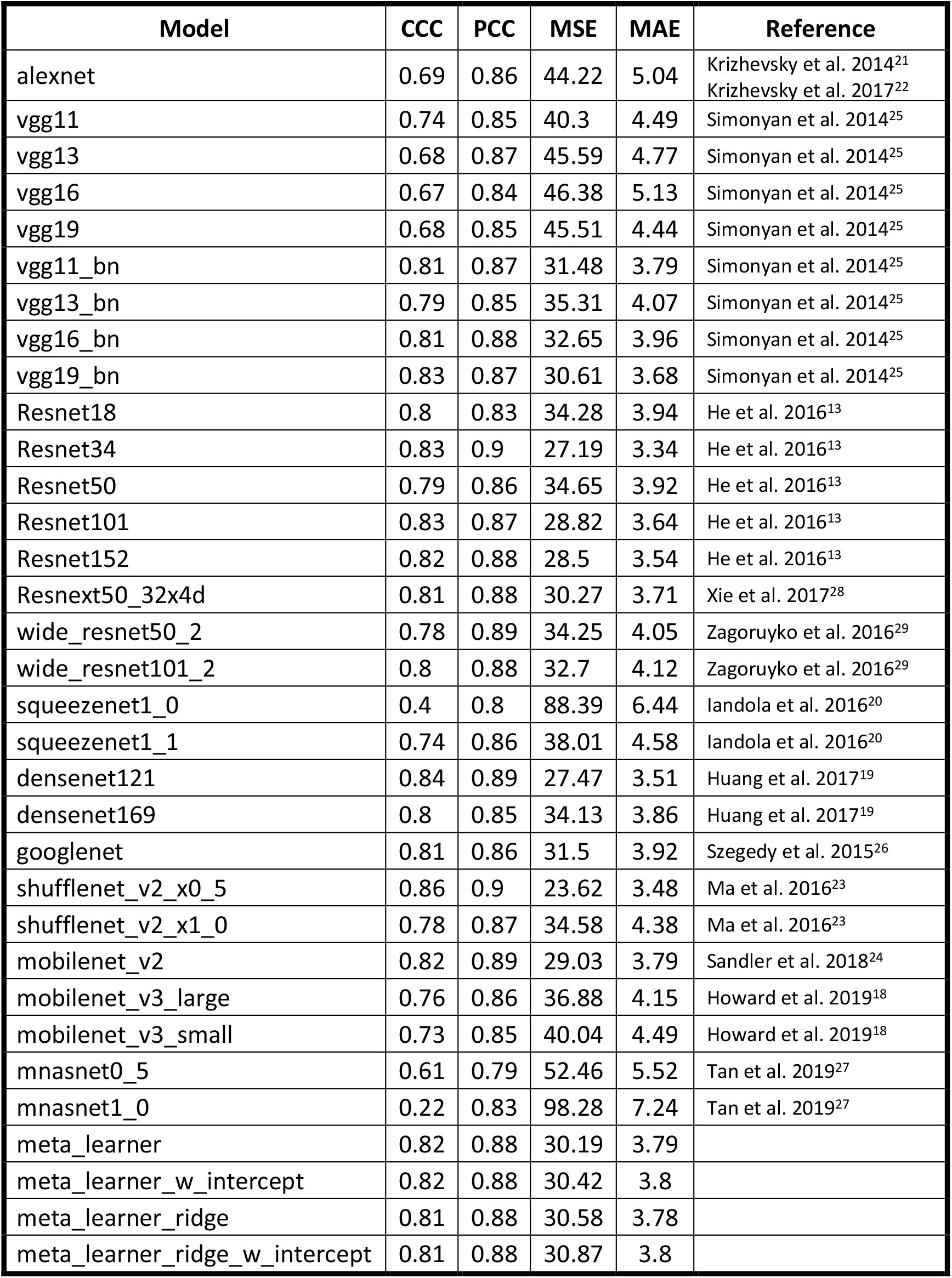
model list of all base and meta learners and their performance on the test dataset.^13, 18, 19, 20, 21, 22, 23, 24, 25, 26, 27, 28, 29^

| Model | CCC | PCC | MSE | MAE | Reference |
| --- | --- | --- | --- | --- | --- |
| alexnet | 0.69 | 0.86 | 44.22 | 5.04 | Krizhevsky et al. 2014<sup>21</sup><br>Krizhevsky et al. 2017<sup>22</sup> |
| vgg11 | 0.74 | 0.85 | 40.3 | 4.49 | Simonyan et al. 2014<sup>25</sup> |
| vgg13 | 0.68 | 0.87 | 45.59 | 4.77 | Simonyan et al. 2014<sup>25</sup> |
| vgg16 | 0.67 | 0.84 | 46.38 | 5.13 | Simonyan et al. 2014<sup>25</sup> |
| vgg19 | 0.68 | 0.85 | 45.51 | 4.44 | Simonyan et al. 2014<sup>25</sup> |
| vgg11_bn | 0.81 | 0.87 | 31.48 | 3.79 | Simonyan et al. 2014<sup>25</sup> |
| vgg13_bn | 0.79 | 0.85 | 35.31 | 4.07 | Simonyan et al. 2014<sup>25</sup> |
| vgg16_bn | 0.81 | 0.88 | 32.65 | 3.96 | Simonyan et al. 2014<sup>25</sup> |
| vgg19_bn | 0.83 | 0.87 | 30.61 | 3.68 | Simonyan et al. 2014<sup>25</sup> |
| Resnet18 | 0.8 | 0.83 | 34.28 | 3.94 | He et al. 2016<sup>13</sup> |
| Resnet34 | 0.83 | 0.9 | 27.19 | 3.34 | He et al. 2016<sup>13</sup> |
| Resnet50 | 0.79 | 0.86 | 34.65 | 3.92 | He et al. 2016<sup>13</sup> |
| Resnet101 | 0.83 | 0.87 | 28.82 | 3.64 | He et al. 2016<sup>13</sup> |
| Resnet152 | 0.82 | 0.88 | 28.5 | 3.54 | He et al. 2016<sup>13</sup> |
| Resnext50_32x4d | 0.81 | 0.88 | 30.27 | 3.71 | Xie et al. 2017<sup>28</sup> |
| wide_resnet50_2 | 0.78 | 0.89 | 34.25 | 4.05 | Zagoruyko et al. 2016<sup>29</sup> |
| wide_resnet101_2 | 0.8 | 0.88 | 32.7 | 4.12 | Zagoruyko et al. 2016<sup>29</sup> |
| squeezenet1_0 | 0.4 | 0.8 | 88.39 | 6.44 | landola et al. 2016<sup>20</sup> |
| squeezenet1_1 | 0.74 | 0.86 | 38.01 | 4.58 | landola et al. 2016<sup>20</sup> |
| densenet121 | 0.84 | 0.89 | 27.47 | 3.51 | Huang et al. 2017<sup>19</sup> |
| densenet169 | 0.8 | 0.85 | 34.13 | 3.86 | Huang et al. 2017<sup>19</sup> |
| googlenet | 0.81 | 0.86 | 31.5 | 3.92 | Szegedy et al. 2015<sup>26</sup> |
| shufflenet_v2_x0_5 | 0.86 | 0.9 | 23.62 | 3.48 | Ma et al. 2016<sup>23</sup> |
| shufflenet_v2_x1_0 | 0.78 | 0.87 | 34.58 | 4.38 | Ma et al. 2016<sup>23</sup> |
| mobilenet_v2 | 0.82 | 0.89 | 29.03 | 3.79 | Sandler et al. 2018<sup>24</sup> |
| mobilenet_v3_large | 0.76 | 0.86 | 36.88 | 4.15 | Howard et al. 2019<sup>18</sup> |
| mobilenet_v3_small | 0.73 | 0.85 | 40.04 | 4.49 | Howard et al. 2019<sup>18</sup> |
| mnasnet0_5 | 0.61 | 0.79 | 52.46 | 5.52 | Tan et al. 2019<sup>27</sup> |
| mnasnet1_0 | 0.22 | 0.83 | 98.28 | 7.24 | Tan et al. 2019<sup>27</sup> |
| meta_learner | 0.82 | 0.88 | 30.19 | 3.79 |  |
| meta_learner_w_intercept | 0.82 | 0.88 | 30.42 | 3.8 |  |
| meta_learner_ridge | 0.81 | 0.88 | 30.58 | 3.78 |  |
| meta_learner_ridge_w_intercept | 0.81 | 0.88 | 30.87 | 3.8 |  |

## Disclosure

This manuscript was sponsored by AbbVie. AbbVie contributed to the design, research, and interpretation of data, writing, reviewing, and approving the publication. All authors are employees of AbbVie Inc. and may own AbbVie stock.

## Code Availability Statement

The code used to generate the models in this paper is available upon request.

## Author Contribution Statements

YX, SZ, and LW designed this study and wrote the first manuscript draft. YX, SZ, and DW performed the analyses. YX, SZ, SA, FR, DW, MC, and LW contributed to the writing, interpretation of the content, and editing of the manuscript, revising it critically for important intellectual content. All authors take accountability for ensuring that questions related to the accuracy or integrity of any part of the work are appropriately investigated and resolved.

## Notes

### Clinical Protocols

https://clinicaltrials.gov/ct2/show/NCT02684357

### Funding Statement

This study was funded by AbbVie Inc.

### Author Declarations

This study utilizes data from the UltIMMA-2 trial, which was done in accordance with the study protocol, International Council for Harmonisation of Technical Requirements for Pharmaceuticals for Human Use (ICH) guidelines, applicable local regulations, and Good Clinical Practice guidelines governing clinical study conduct, and ethical principles outlined in the Declaration of Helsinki. All study-related documents (including the study protocols) were approved by an institutional review board or independent ethics committee at each study site, and all patients provided written informed consent before participation. More details can be found on ClinicalTrials.gov with identifier NCT02684357

